# Sentinel surveillance of SARS-CoV-2 in wastewater anticipates the occurrence of COVID-19 cases

**DOI:** 10.1101/2020.06.13.20129627

**Authors:** Gemma Chavarria-Miró, Eduard Anfruns-Estrada, Susana Guix, Miquel Paraira, Belén Galofré, Gloria Sánchez, Rosa M. Pintó, Albert Bosch

**Affiliations:** Enteric Virus laboratory, Department of Genetics, Microbiology and Statistics, Section of Microbiology, Virology and Biotechnology, School of Biology, and Institute of Nutrition and Food Safety, University of Barcelona, Barcelona, Spain; University of Barcelona, Spain; Aigües de Barcelona, Barcelona, Spain; Department of Preservation and Food Safety Technologies, Institute of Agrochemistry and Food Technology, IATA-CSIC, Paterna, Valencia, Spain

**Keywords:** SARS-CoV-2, COVID-19, epidemiology, surveillance, early warning, sewage, wastewater

## Abstract

SARS-CoV-2 was detected in Barcelona sewage long before the declaration of the first COVID-19 case, indicating that the infection was present in the population before the first imported case was reported. Sentinel surveillance of SARS-CoV-2 in wastewater would enable adoption of immediate measures in the event of future COVID-19 waves.

**Article Summary Line:** SARS-CoV-2 genomes occur in sewage long before the declaration of COVID-19 cases among the population.

---

In early December 2019, COVID-19 originated in Wuhan, China, and reached thereafter many parts of the world, including Europe, where the first case was reported in France in late-January 2020 (1). However, evidence points to the occurrence of cases in France already in late 2019.

Despite COVID-19 is a respiratory disease, large amounts of coronavirus genomes are shed in the feces (2, 3) and ultimately reach wastewater (4, 5). SARS-CoV-2 surveillance in sewage may be considered a sensitive tool to monitor the spread of the virus among the population. However, there is no epidemiological evidence that sewage could be a transmission route for SARS-CoV-2, through contamination of bathing areas or irrigation waters, because very few studies report culture of infectious virus from stool (6). Even in respiratory samples, infectivity is only associated with very high genome copy numbers (7, 8) that very unlikely are found in wastewater.

To date, Spain ranks in fourth place in absolute number of cases, and almost topping the list in number of cases and deaths per 1 million inhabitants, being Barcelona the second most affected area. The first case in Barcelona was reported in February 25, 2020 and the total number of cases at the end of May 2020 was over 17,500 (https://salutweb.gencat.cat/ca/inici/nota-premsa/index.html?id=385948#googtrans(ca|en)).

## The Study

In order to elucidate the evolution of COVID-19 in Barcelona, 24-h composite raw sewage samples from two large wastewater treatment plants (WWTP1 and WWTP2) were weekly analyzed for the presence of SARS-CoV-2 from April 13, in the peak of the epidemics, to May 25. In addition, for WWTP2, frozen archival samples from 2018 (January-March), 2019 (January, March, September-December) and 2020 (January-March) were also assayed.

Eight hundred-milliliter samples of wastewater were concentrated through precipitation with 20% polyethylene-glycol 6000 and resuspended in 3 mL of PBS, pH 7.4 (9). Nucleic acid extraction was performed from 1mL of the concentrate and eluted in 50 µL using the NucliSENS® miniMAG® extraction system (bioMérieux).

Five one-step RT-qPCR assays (RNA UltraSense(tm) One-Step Quantitative RT-PCR System, Invitrogen, Life Technologies) targeting the RNA-dependent RNA polymerase (RdRp) gene, IP2 and IP4 fragments, from Institute Pasteur, Paris (Institut Pasteur. Protocol: Real-time RT-PCR assays for the detection of SARS-CoV-2. 2020 https://www.who.int/docs/default-source/coronaviruse/realtime-rt-pcr-assays-for-the-detection-of-sars-cov-2-institut-pasteur-paris.pdf?sfvrsn=3662fcb6_2), the envelope protein (E) gene, E fragment, from Charité, Berlin (10), and the nucleoprotein (N), N1 and N2 fragments, from CDC, Atlanta (Centers for Disease Control and Prevention. CDC 2019-Novel Coronavirus (2019-nCoV) Real-Time RT-PCR Diagnostic Panel. 2020 https://www.fda.gov/media/134922/download). The standard curve were constructed using the Twist Synthetic SARS-CoV-2 RNA Control 2 (MN908947.3) (Twist Bioscience). Technical details are included in the Appendix.

In WWTP1, SARS-CoV-2 genome copy numbers progressively decreased from April 13 to May 18. This decrease was observed employing IP2 and IP4 targets (Figure 1, panel A), and confirmed with E, and N1 and N2 targets (Figure 1, panels B and C). On May 18, genomes copies were below the theoretical detection limit, although residual levels could be again detected on May 25 employing the N1 target. SARS-CoV-2 genome copy levels in wastewater clearly matched the estimated cumulative population shedding virus in stool (Figure 1, panel K, L, M).

**Figure 1.**
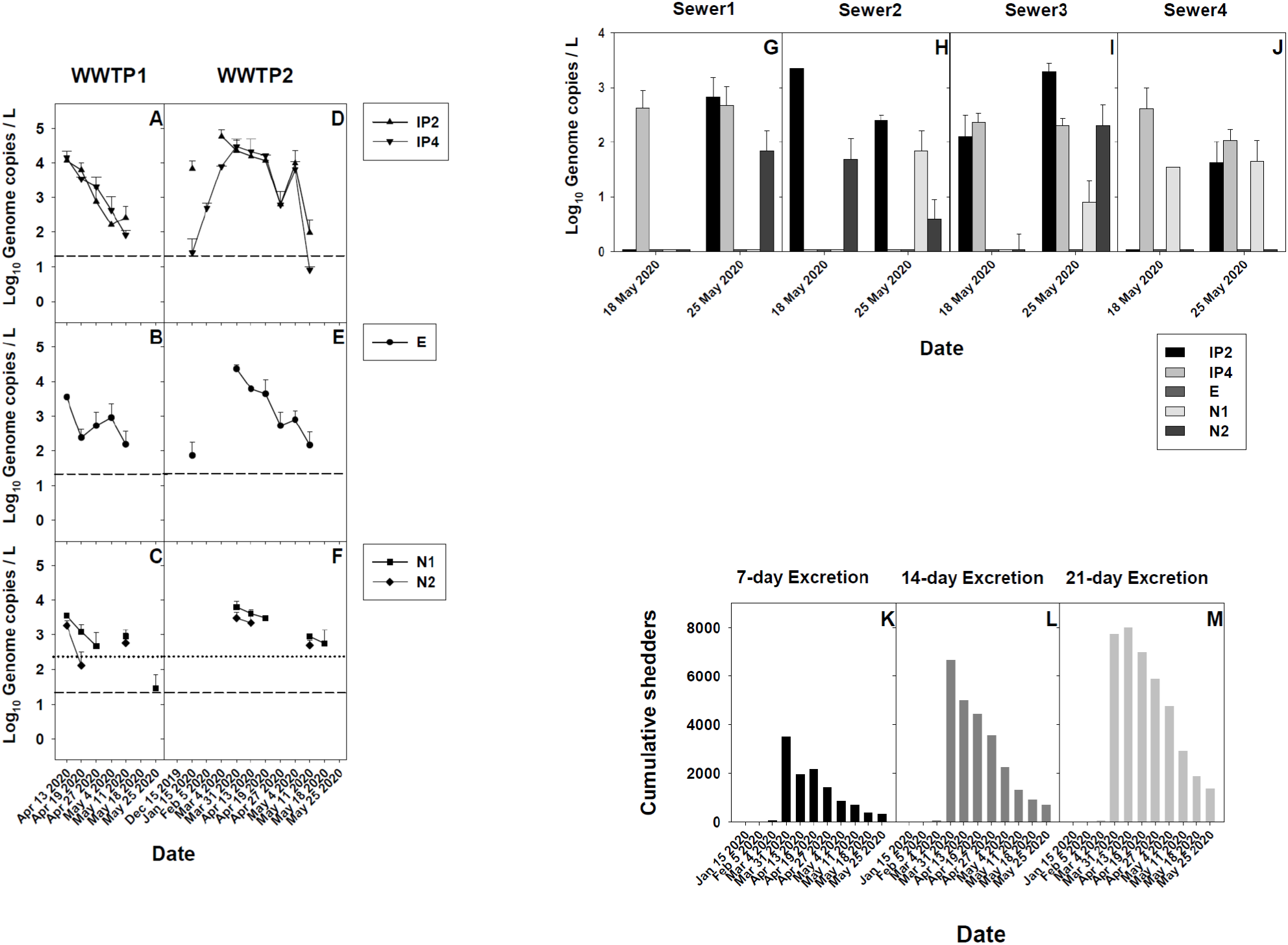
SARS-CoV-2 in Barcelona sewage and its association with COVID-19 cases. Panels A-F: SARS-CoV-2 genome copy levels in two WWTP, detected with targets IP2 and IP4 (A, D), E (B, E) and N1 and N2 (C, F). Each point represents the mean and standard deviation of 2-4 replicas of RT-qPCR assays. Absence of values at a given date means genome copy numbers below the theoretical limit of detection (dashed line for IP2, IP4, E and N2, and dotted line for N1), with the exception of N2 target on April 27 and May 4 for which aliquots to assay were no longer available. Some samples showed positivity below the theoretical limit of detection (WWTP1: N1, May 25 2020; WWTP2: IP4, May 11 2020). Panels G-J: SARS-CoV-2 genome copy levels in four urban sewers, detected with targets IP2, IP4, E, N1 and N2. Panels K-M: Cumulated SARS-CoV-2 shedders figured estimating fecal excretion periods of 7 days (K), 14 days (L) and 21 days (M) prior to case reporting.

For WWTP2, samples from December 2019 to May 2020 were available, which opened the possibility to better analyze the dynamics of genome copy numbers in sewage. These numbers, determined with IP2 and IP4 targets, followed a curve peaking between March 4 and May 4 (Figure 1, panel D). In the middle of the peak, an abrupt fall in genome copies was observed on April 27, likely due to a heavy rainfall event occurring during April 19-21, with daily pluviosity averages of 65 mm. These rainwaters are collected in storm water reservoirs of 375,500 m^3^ that gradually drain into sewers, adding a considerable dilution factor to viruses in wastewater. This dilution effect was not observed in WWTP1 samples, since no large storm water reservoirs affect its sewerage network. A progressive decline in genome copy numbers in WWTP2 parallels the diminution in the number of estimated virus shedders in the community (Figure 1, panel K, L, M), as observed in WWTP1. Nevertheless, on May 18, genomes could still be detected employing the N1 target although completely disappeared on May 25 (Figure 1, panel F).

Unexpectedly, analysis of archival samples revealed the increasing occurrence of SARS-CoV-2 genomes in samples from January 15 to March 4, 2020 (Figure 1, panels D and E). Of note, SARS-CoV-2 was detected in sewage 41 days (January 15) before the declaration of the first COVID-19 case (February 25), clearly evidencing the validity of wastewater surveillance to anticipate cases in the population. This SARS-CoV-2 early detection in sewage supports the idea that COVID-19 cases may have been present in the population before the first imported case was reported. COVID-19 carriers may have been misclassified as influenza diagnoses in primary care, boosting community transmission before public health measures were taken (11). Additionally, there is a significant proportion of asymptomatic carriers that shed SARS-CoV-2 and contribute to the virus spread (12).

Despite the apparent disappearance of SARS-CoV-2 in WWTP1 and WWTP2 around May 18-25, the analysis of grab samples, collected 8-9 AM, from four urban sewers revealed the occurrence of virus genomes (Figure 1, panels G-J). Sewer1 drains into WWTP1, while sewer2, sewer3 and sewer4 drain into WWTP2. A higher dilution factor applies in the WWTPs than in the sewers, which together with differences applying between grab and composite samples, and bowel habits (13), could explain why samples from the former came out to be negative for the virus, while genome copies could still be detected in the sewer samples. Sewer analysis may provide most relevant information on the localization of areas where COVID-19 cases reappear, enabling immediate response to prevent spread of the outbreak. Nevertheless, it represents a more laborious and costly approach that surveillance through WWTP monitoring.

Most COVID-19 cases show mild influenza-like symptoms (14) and it has been suggested that some uncharacterized influenza cases may have masked COVID-19 cases in the 2019-2020 season (11). This possibility prompted us to analyze some archival WWTP samples from January 2018 to December 2019 (Figure 2). All samples came out to be negative for the presence of SARS-CoV-2 genomes with the exception of March 12, 2019, in which both IP2 and IP4 target assays were positive. This striking finding indicates circulation of the virus in Barcelona long before the report of any COVID-19 case worldwide. Barcelona is a business and commerce hub, as well as a popular venue for massive events, gathering visitors from many parts of the world. It is nevertheless likely that similar situations may have occurred in several other parts of the world, with circulation of unnoticed COVID-19 cases in the community.

**Figure 2.**
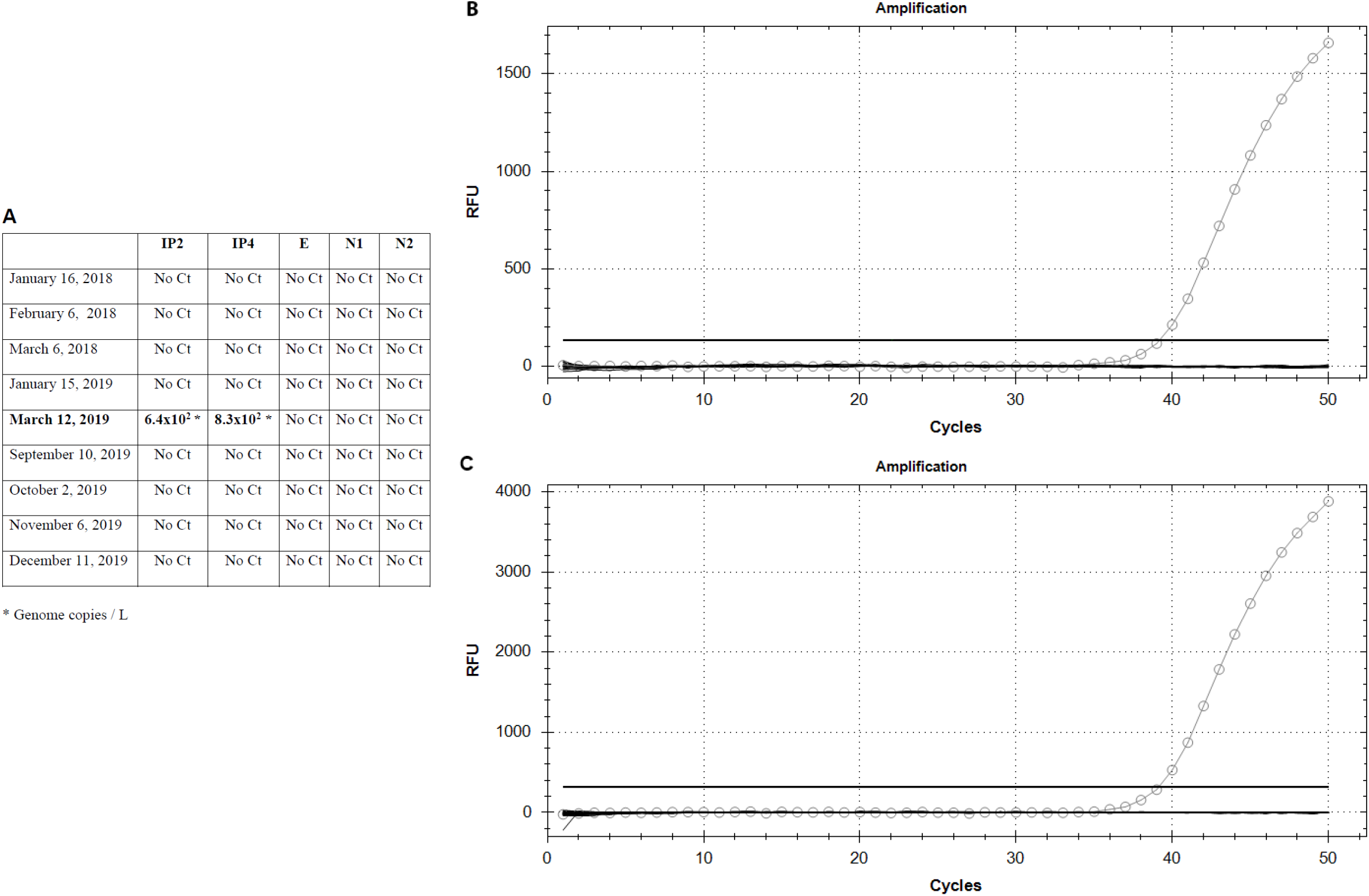
SARS-CoV-2 detection in archival samples collected from January 2018 to December 2019 at WWTP2. A. SARS-CoV-2 genome detection with targets IP2 and IP4, E and N1 and N2. B. Real-time RT-PCR amplification plots of IP2 and IP4 targets in the March 12, 2019 sample.

## Conclusions

A significant proportion of undiagnosed and asymptomatic carriers shed SARS-CoV-2 in stool. Wastewater-based epidemiology is an early warning tool of the circulation of the virus in the population, including symptomatic and asymptomatic shedders. In the specific case of Barcelona, awareness of SARS-CoV-2 spread with over one-month anticipation would have enabled a better response to the epidemics. The enormous burden in morbidity and mortality of COVID-19 calls for sentinel surveillance of SARS-CoV-2 in wastewater to enable rapid mitigation measures in the likely event of a future pandemic wave of the infection.

## Data Availability

All raw data will be provided upon request

## Acknowledgements

This research was supported in part by the REVEAL project, funded by SUEZ Spain. We thank our colleagues from Aigües de Barcelona, Cetaqua and Labaqua, who provided insight and expertise for the research. We are indebted with the Metropolitan Area of Barcelona (AMB) for a fruitful collaboration. We are grateful to M. Jané-Checa and A. Martínez-Mateo of the Public Health Agency of Catalonia (ASPCAT) for providing epidemiological data and useful discussion.

